# Change in mental health trends in Scotland, UK pre- and post-COVID-19

**DOI:** 10.1101/2024.12.16.24319095

**Authors:** Elizabeth A. Cooke, Agnieszka Lemanska, Spencer A. Thomas

**Author notes:** Corresponding Author: Dr Elizabeth Cooke, National Physical Laboratory, Hampton Road, TW11 0LW Middlesex, UK.

## Abstract

The COVID-19 pandemic and national lockdowns have had profound impacts on population mental health in Scotland. In this study, we examine the impact on the relationship between the number of patients receiving in-patient care for mental health related diagnoses and the number of patients on waiting lists for mental health treatment. Prior to March 2020, increased waiting lists did not translate into an increase in hospital admissions. Post-March 2020, however, we found that more patients on waiting lists corresponded to more patients being hospitalised. Our analysis indicates that the relationship between waiting lists and in-patient treatment changed during the COVID-19 pandemic, and this may be contributing to additional pressures on hospitals and mental health services in Scotland.

## 1. Introduction

In response to the COVID-19 pandemic the UK Government issued a national lockdown in March 2020. Both the pandemic and national lockdowns have had profound impacts on population mental health due to lockdown isolation [1], increased levels of anxiety [2,3] and limited access to healthcare services and face-to-face consultations [4].

Previously, hospital admissions for mental health related diagnoses in England and Scotland have been shown to be falling over time, and the average length of hospital stay has decreased since 2014 [5,6]. However, the number of readmissions has been increasing since 2014 in Scotland [5], and the COVID-19 pandemic increased waiting times for mental health treatment for both adults and children and adolescents [7,8].

Recently, the agency for health and wellbeing in Scotland, Public Health Scotland (PHS) have highlighted the need to improve the mental wellbeing of the population using data [9] and the Scottish government have made improving mental health support a priority [10]. Increased attention on mental health may lead to increased demand on services, but resources and capacity may not scale at the same rate. Moreover, the full and long term impact of the COVID-19 pandemic on access to services may not have been fully realised. In this study, we investigated whether the number of patients receiving in-patient care for mental health related diagnoses in Scotland correlates with the number of patients on waiting lists for mental health therapies, and the effect of the COVID-19 pandemic on that relationship.

## 2. Methods

We used PHS national registers of patient data showing the number of patients on waiting lists for mental health treatment [11] and admitted to hospital for mental health related diagnoses [12]. The data ranged from financial years (April–March) 2014/15 to 2022/23 (2023/24 for waiting list data). Diagnoses were coded using the ICD-10 coding system [13], using F00–F99 to classify mental health conditions. Population data for Scotland was obtained from the Scottish Health and Social Care Open Data [14].

The waiting list data showed the total number of patients on waiting lists for mental health treatment prior to their initial consultation each month. These data were combined into financial years and matched by year to the in-patient stay data. For each year, the total number of patients on waiting lists was compared to the number of patients in hospital for mental health-related diagnoses. The number of in-patients was categorised by the length of stay: < 1 day, 1 to 7 days, 8 to 28 days, 29 days to 6 months, and > 6 months. We presented the rates of in-patient stays per 100,000 population, and their relationship with numbers of patients waiting for mental health treatment. We compared the trends from pre- and post-March 2020, which corresponds to the start of the COVID-19 national lockdown in the UK.

## 3. Results

Figure 1 shows the number of in-patient stays, normalised by population size, and their relationship with patients waiting for mental health treatment pre- and post-lockdown. Prior to the national lockdown, the numbers of in-patients per 100,000 population were decreasing, or remaining flat, while numbers of patients waiting for treatment were increasing. Following the national lockdown, this relationship changed dramatically with numbers of patients waiting for treatment and numbers of hospital admissions increasing for stays up to 28 days, and decreasing for stays of 29 days and longer. Overall, post-lockdown, shorter stays exhibited an increasing number of stays for increasing number of patients waiting, with the inverse recorded for the pre-lockdown trend. The number of stays, for both shorter and longer durations, exhibited a steeper dependency on number of patients waiting post-lockdown than prior.

**Figure 1.**
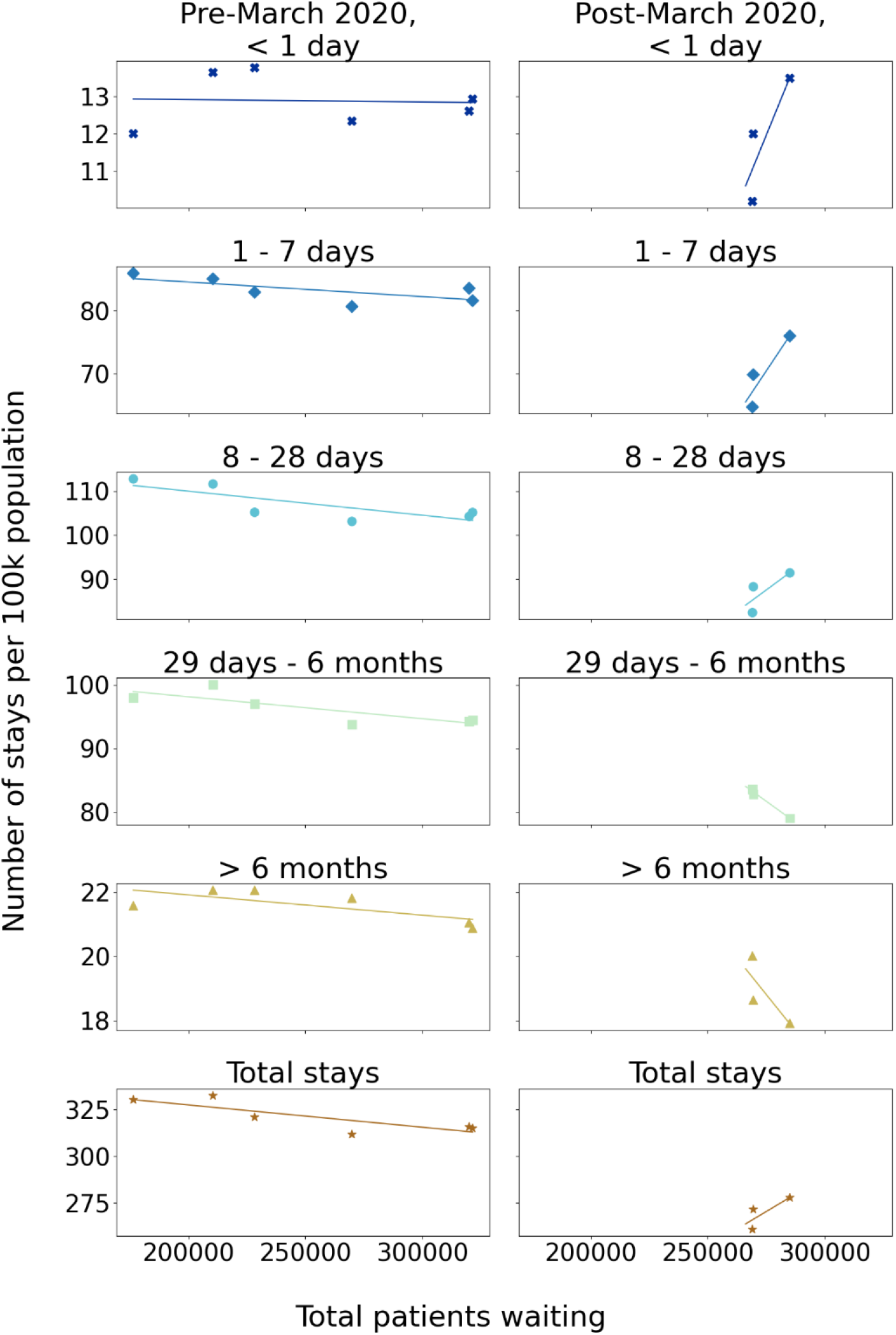
The number of in-patient stays, normalised by population size (y-axes), and their relationship with patients waiting for mental health treatment x-axes), pre- (left panel) and post- (right panel) the start of the national lockdown due to the COVID-19 pandemic. Rows show different lengths of stay times, with the bottom graphs showing the relationship for all lengths of stays together.

Figure 2 shows that, for the period from 2014/15 to 2019/20, waiting lists for mental health treatment grew from 176,000 to 321,000 patients then decreased to 285,000 in 2020/21 and further fell to 269,000 in 2023/24. From the increase in patients waiting from 2014/15 to 2019/20 we can infer that the decrease in number of stays in the left column of Figure 1 was a chronological trend. The drop in total patients on waiting lists following March 2020 has not recovered and remained flat up to March 2024.

**Figure 2.**
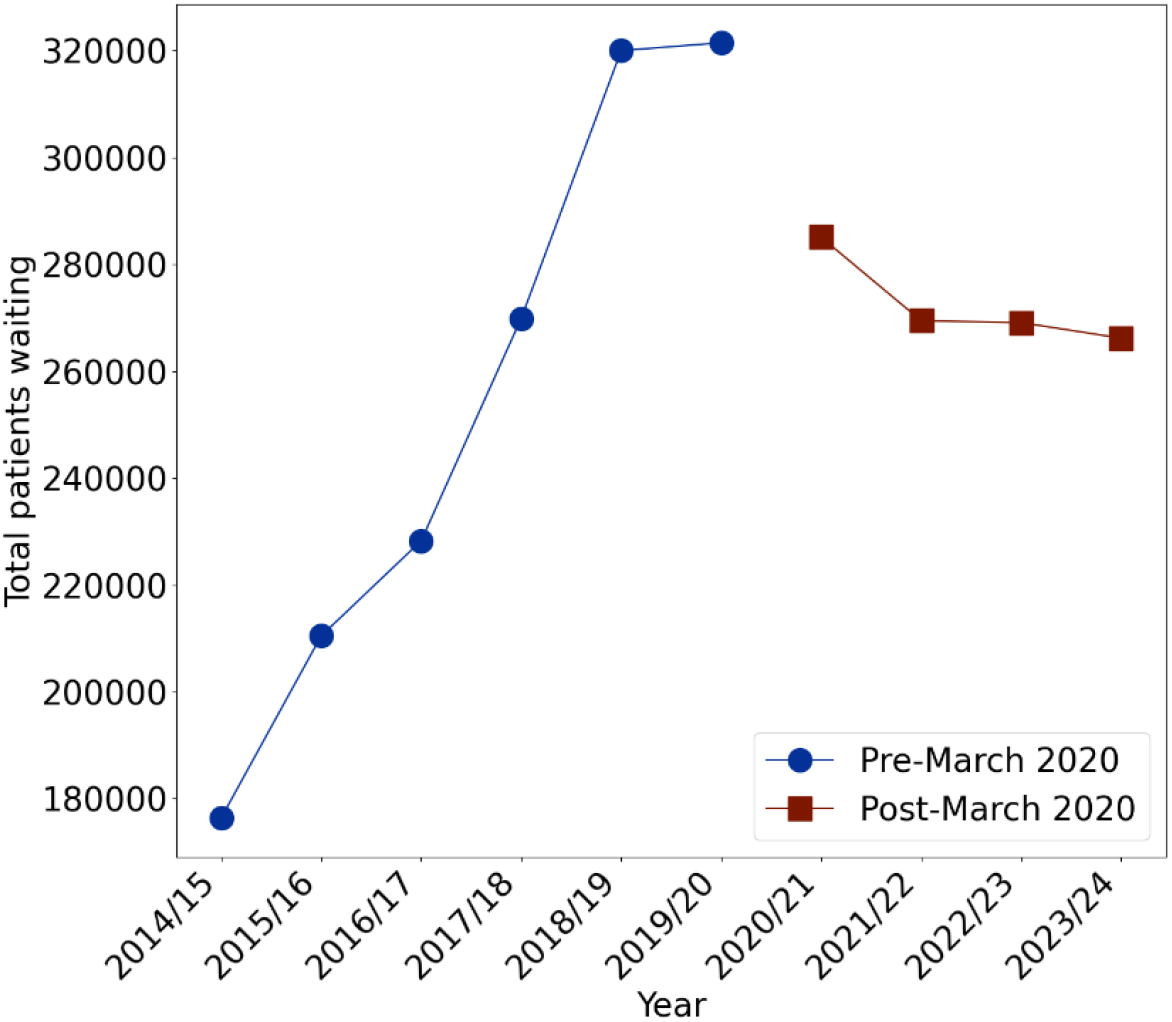
Number of patients waiting for psychological therapies over time. Before March 2020, waiting lists were increasing. During 2020/21, the number of patients on waiting lists dropped dramatically and has flattened since.

## 4. Discussion and conclusions

The COVID-19 pandemic was linked to a change in the relationship between numbers of in-patient stays and number of patients on mental health treatment waiting lists. This effect was recorded for all lengths of stay. The impact of the COVID-19 pandemic and subsequent national lockdowns may have changed the relationship between patients waiting for mental health treatment and the number of patients requiring hospitalisation for treatment. In particular, more patients on waiting lists appeared to relate to more patients being hospitalised for short (≤ 28 days) in-patient stays. We note, however, that there was insufficient data to quantitatively assess the post-lockdown trends and more data is required to confirm these observations. Prior to March 2020, increased waiting lists did not translate into an increase in hospital admissions. This may have been due to more care occurring outside of secondary care, which is now being handled by hospitals.

Our analysis indicates that the relationship between waiting lists and in-patient treatment has changed, which may put additional pressure on hospitals and mental health services in Scotland. Further work is required to better understand the impact of the COVID-19 pandemic and subsequent lockdowns and whether these patterns will continue in future years. Efforts to lower waiting times for mental health treatment are needed if Scotland is to achieve its goal of improving the mental health and wellbeing of the population.

## Data Availability

All data produced are available online at https://www.opendata.nhs.scot/dataset

## Acknowledgements

This work was funded by the UK Government’s Department for Science, Innovation & Technology (DSIT) through the UK’s National Measurement System programmes.

